# Understanding Barriers toward Interventions for Childhood Obesity in Minority Communities: A Rapid Review

**DOI:** 10.1101/2022.03.29.22273114

**Authors:** Kenya M. Colvin

## Abstract

**Background:** It is well documented that racial/ethnic minorities have higher attrition rates from pediatric weight manage programs than their white counterparts. This review aimed to identify barriers facing minority families when seeking to complete weight management programs for their children to develop strategies to keep these families engaged. Maintaining engagement through program completion could improve health outcomes and overall quality of life for minority children.

**Methods:** Eligible studies were identified through searching PubMed, PsychINFO and CENTRAL databases. Articles had to focus on reasons for attrition from pediatric weight management among minority groups to be included. We extracted data on attrition from each study and narratively summarized the results.

**Results:** From the 302 articles screened, five met the inclusion criteria. Four of the five studies predicted attrition factors from attendance or past survey data. Only one study surveyed parents to identify potential barriers. Among these studies, most sociodemographic factors analyzed had no significant effect on early dropout. Program structure, logistical barriers, and parental negative self and child body image were all identified as predictors of attrition.

**Conclusion:** More studies need to directly investigate why minority families discontinue weight management treatments early. Directly interviewing or surveying parents to ascertain their concerns can lead to the development of programs that retain and better meet the needs of these groups.

## Introduction

In the midst of the global COVID-19 pandemic, obesity remains one of the most pressing public health concerns of our time^1–3^. In the last twenty years, global rates of obesity have risen to alarming levels, especially among children^1,4^. Lifestyle alterations and restrictions ensuing from the COVID-19 pandemic compounded with pre-existing factors further increased the worldwide prevalence of childhood obesity^5,6^. Obesity is defined by the World Health Organization as excessive or abnormal fat accumulation that may negatively impact overall health^7^. Body mass index (BMI) is routinely used as a diagnostic measure to assess obesity or overweight status for everyone over age two^8^. The Centers for Disease Control and Prevention (CDC) defines childhood obesity as having a BMI ≥ 95^th^ percentile for their age and sex^9^. Overweight is defined as BMI ≥ 85^th^ percentile to less than the 95^th^%ile for age and sex^9^. The U.S. 2017-2018 National Health and Nutrition Examination Survey (NHANES) found that the prevalence of obesity among U.S. children and adolescents (2-19 years of age) was 19.3%^10^. The prevalence of overweight children and adolescents in this time frame was 16.1%^10^. Despite the widespread disease burden of obesity across all racial and ethnic groups within the United States, minority groups (Black/African America, Hispanic/Latinx, Native Hawaiian, Pacific Islander, Native American/Alaskan Native) are disproportionately affected^11,12^. Black and Hispanic communities bear the greatest impact of this inequity^13^. From the NHANES study, Non-Hispanic Asians had the lowest obesity prevalence (8.7%) while Non-Hispanic Blacks and Hispanics had an obesity prevalence of 24.2% and 25.6% respectively, compared to 16.1% among their non-Hispanic White (NHW) counterparts^10^. A variety of factors, including socioeconomic status, cultural differences, prenatal behaviors, and environment, have been linked to driving this disparity^11–13^.

Obesity can lead to long term health conditions such as type 2 diabetes, dyslipidemia, or coronary artery disease^14,15^. Intervening in childhood is critical to reducing the likelihood of adulthood obesity and chronic disease^16^. Current prevention and intervention methods for children include nutrition counselling and increasing access to healthy foods; weight management programs focused on increasing physical activity; behavioral therapy; pharmacologic therapy; and bariatric surgery^13^. Combined family-based programs that address nutrition, behavior and physical activity are also beneficial^17^. Community and hospital based weight management programs are widely available, but are dependent on continued family participation for success^17^. Attrition from these programs is persistently around 30-40%^18^. However, premature program departure can be as high as 75-83%^17–19^. Minorities and families of low socioeconomic status have the highest dropout rates from obesity management programs, despite having the greatest treatment need^18,20^. Understanding why families leave these programs early is essential to reducing obesity and overall health status among members of these communities^18^. Several reviews have explored attrition from pediatric weight management programs^17,18,21,22^, but very few in the context of Black/African American and Hispanic/Latinx communities^20^. Ligthart et al. examine the interrelationship between ethnicity, socioeconomic status, and pediatric obesity management programs^20^. The study concluded that ethnic minorities, especially African Americans, have higher dropout rates, but did not investigate why these groups were more likely to discontinue treatment^20^. They acknowledge in their limitations that other variables likely play a role but were not able to be incorporated in their study^20^.

The aim of this rapid review was to systematically search and analyze the literature to identify barriers faced by Black and Hispanic families in accessing or continuing weight management programs to provide strategies to improve patient retention and outcomes among these groups.

## Methods

### Search Strategy

A systemic approach was taken to explore literature in PubMed, CENTRAL, and PsychINFO with no time constraints. Searches were conducted in January and February 2022. Google Scholar was also used to gather relevant sources to help define terms and construct search methods. A preliminary examination of systematic reviews on attrition in pediatric weight management was conducted to identify narrower search terms. An experienced informationist aided in the development of the search strategy. A sequential search strategy was employed for all three databases. Search terms were tailored to each database, but were all related to one of the following categories: minority groups, childhood/adolescence, obesity, and treatment compliance. “Cited by” and “similar article” listings from relevant search results were also reviewed to identify additional studies. Search strategies used are included in Supplemental Table 1. Zotero 5.0 for Windows was used to manage and export citations.

### Study Selection

Studies that met the following criteria were eligible for inclusion: (a) age of population examined fell between 2 -21 years old, (b) investigated reasons for low compliance or attrition from obesity treatment programs, (c) children in study were diagnosed with obesity/overweight, (d) at least 70% of participants were members of a racial/ethnic minority group, (e) written in English, and (f) was a primary research study (e.g. RCT, cross-sectional, cohort, or qualitative). Article titles and abstracts were first evaluated to determine if they fit the inclusion criteria. If the initial screenings were passed, the full article was read to finalize determination of eligibility. Studies that included adults, did not focus on minority communities, or only analyzed treatment outcomes without regard for compliance were excluded. Failure to pass both title and abstract screening also resulted in exclusion. Figure 1 shows the results from the previously described search strategy in accordance with the Preferred Reporting Items for Systematic Reviews and Meta Analyses: The PRISMA Statement^23^.

**Fig. 1.**
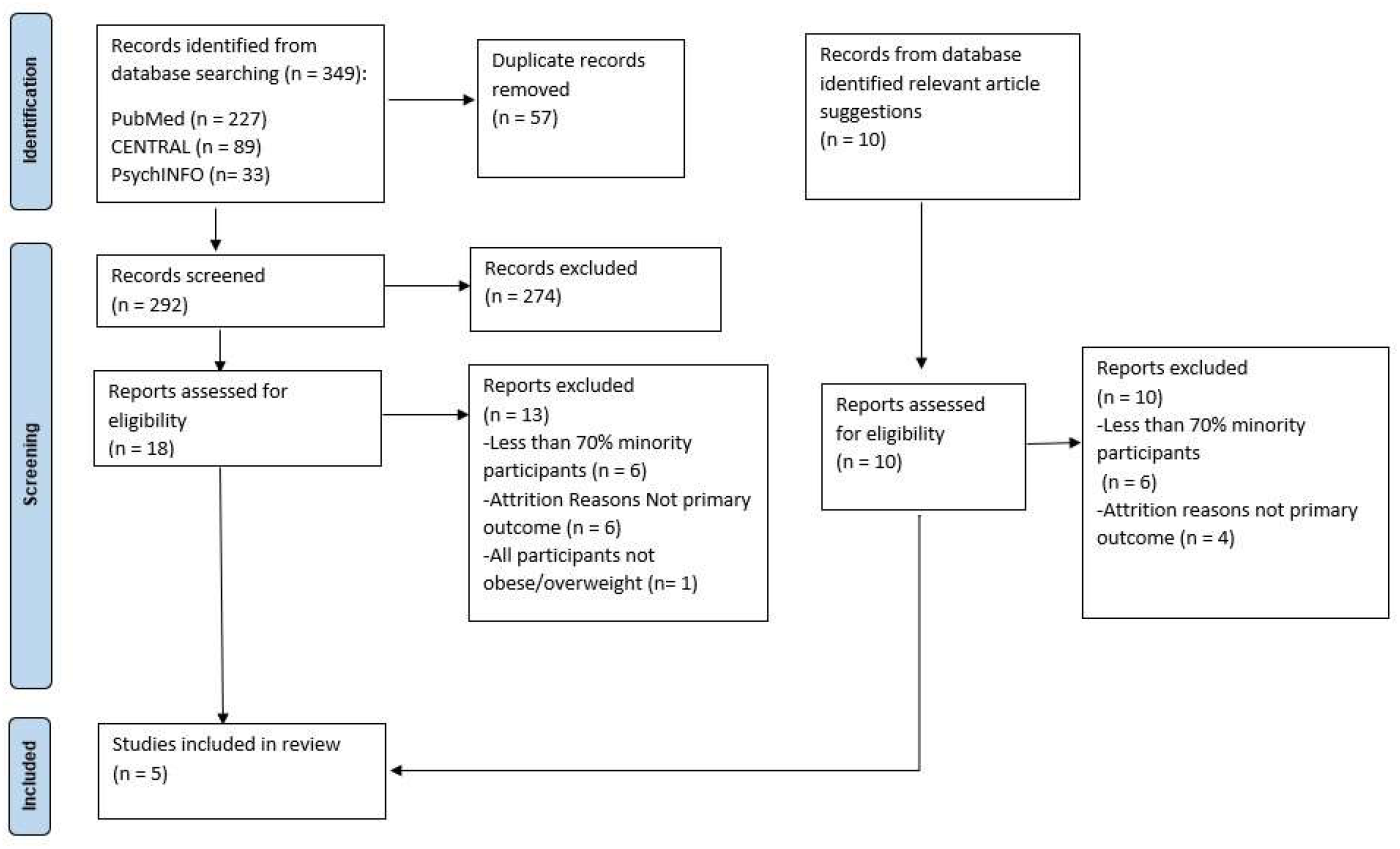
Prisma flow diagram of study selection process with reasons for exclusion.

### Data extraction and Analysis

Data was extracted from non-RCTs and RCTs using data extraction forms adapted from Cochrane templates^24^. We extracted only data, measurements and statistical analyses related to attrition outcomes. Data items extracted included: study setting; aims; sociodemographics; intervention program description; findings related to attrition; author recommendations; and key limitations. Descriptive analysis of non-RCTs and RCTs were performed separately and then integrated. All data were summarized narratively.

### Risk of bias assessment

Quality assessments were performed for all included studies using Cochrane’s risk of bias tools. For the RCT the revised tool to assess risk of bias in randomized trials (RoB 2) for cluster trials was used^25^. For all non-randomized trials, the risk of bias in non-randomized studies - of interventions (ROBINS-I) tool was used^26^. Studies were rated as having low, moderate, serious, or critical risks of bias. Quality ratings were not used as exclusion criteria.

## Results

A total of 302 articles were screened after the removal of duplicates. Among those studies, only 28 studies passed title and abstract screening. Full length articles of all 28 studies were retrieved and only five met all inclusion criteria^27–31^. One report was a cluster randomized controlled trial^30^ and the others were non-RCTs^27–29,31^. The non-RCTs included two retrospective cohort studies^27,28^, one cross-sectional study^29^ and one pre-post study^31^. All of the reports, except for the cross-sectional one, identified possible predictors of attrition from a lifestyle change program. The cross-sectional study^29^ examined parent-reported barriers to completion of a pediatric obesity intervention directed towards parents. Each of the investigations included at least one parent along with their child as participants. Three of the five interventions were family based^28,30,31^,one was parent only^29^, and for one, the activities were primarily child focused but mothers completed the study questionnaire analyzed^27^. Among the reports investigating predictors of attrition^27,28,30,31^, three included both Black and Hispanic participants^27,28,31^, while one examined an intervention for only Hispanic families^30^. For the investigation focused on Hispanic families, the participants represented immigrants from various Latin American countries (39%) and US born Hispanics (61%)^30^.

Attrition rates were described as high for all the lifestyle change programs examined^27–31^, but the rates were not reported in a consistent manner. The programs varied in length and some represented attrition through categorizing different attendance levels. Notably, Germann et al.^28^ reported a 57% 3-month attrition rate for their program, which was higher than other previously published literature on short term programs. Attending less than 50% of sessions ranged from 19-70%^27,29–31^.

**Table 1.** Summary of included studies characteristics and outcomes related to attrition or specified barriers to program completion. Sample size and race/ethnicity represents the children included in study for sources 28, 29,31, and 32. For source 30, sample size and race/ethnicity describe caregivers.

| Reference | Study location<br>(all in USA) | Study design<br>(barriers predicted<br>or measured) | Sample size (N);<br>%race/ethnicity<br>(Black, Hispanic) | Age range<br>(Years) | Barriers/ predicted<br>attrition factors<br>identified | Bias rating<br>(low,<br>moderate,<br>serious, or<br>critical) |
| --- | --- | --- | --- | --- | --- | --- |
| Dhuper et al.<br>(2020)<sup>27</sup> | Brooklyn, New<br>York | Retrospective<br>Cohort<br>(barriers predicted) | 596; 84%, 15% | 2-19 | Negative maternal<br>perceptions of their<br>own health status,<br>body weight, self-<br>image, and image of<br>their child | Moderate |
| Germann et al.<br>(2006)<sup>28</sup> | Chicago, Illinois | Retrospective<br>Cohort<br>(barriers predicted) | 342; 89.2%,<br>6.1% | Mean age,<br>13.0 | Male gender, lack of<br>program orientation<br>session | Moderate |
| Kwitowski et al.<br>(2017)<sup>29</sup> | Richmond, VA | Cross-sectional<br>(barriers measured<br>via survey) | 70; 71.4%, 2.9% | 5-11 | Program design too<br>parent oriented,<br>inadequate child<br>engagement, logistical<br>barriers, difficulty<br>balancing with other<br>responsibilities | Low |
| St. George et al.<br>(2018)<sup>30</sup> | Miami, FL | RCT<br>(barriers predicted) | 140; NA, 100% | Mean age,<br>13.04 | Higher levels of<br>Americanism in<br>parents and higher<br>levels of Hispanicism<br>in adolescents | Low |
| Williams et al.<br>(2010)<sup>31</sup> | Tennessee (city<br>not specified) | Pretest/posttest<br>(barriers predicted) | 155; 73.55%;<br>2.58% | 4-7 | Lower income, high<br>levels of family<br>dysfunction<br>(specifically<br>disengagement | Low |

A consistent finding across all the studies examining predictors of attritions^27,28,30,31^ was that child BMI differences at baseline was not a significant predictor of attrition. Female gender was found as a significant predictor of compliance in one study^28^, all others found gender to have no bearing on attendance rate^27,30,31^. A common trend was that sociodemographic factors such as parental education level, age and income level were not predictive measures. Income level was reported as significant in only one study and this report had a larger inclusion of White participants (20.65%)^31^ compared to the other predictive studies (< 5% overall)^27,28,30^.

One cross-sectional study used both quantitative and qualitative methods to assess self-reported barriers to pediatric weight management program completion^29^. The intervention from this report was a parent only pediatric weight management program. Thirty-seven percent of caregivers cited too much focus on parents and inadequate child involvement as reasons for early program drop-out. Other caregiver-identified barriers included transportation difficulties, inability to find free parking, program distance and time, scheduling issues and balancing other familial priorities. All of the studies agreed that culturally sensitive and minority targeted weight management programs are needed to retain these groups^27–31^.

## Discussion

This rapid review aimed to explore the predicted and expressed barriers faced by minority families toward completing pediatric obesity intervention programs. Weight management attrition literature focused on primarily minority communities is limited, with several of the studies excluded here included less than 50% minority participants^32–38^.

Despite original studies and systematic reviews citing being a minority as a significant predictor of attrition^20,27,31,32,37^, only minimal research has assessed why these groups drop out early^29^. Dhuper et al. acknowledged the importance of directly examining reasons for attrition among minority groups and planned to investigate this in the future^27^.

In contrast to other reports on attrition from pediatric weight reduction programs^32,38^, BMI at baseline was not a significant predictor of early drop out among primarily minority populations^27– 31^. The BMI of enrolled participants among the included studies was comparable, with a large proportion having severe obesity^27–31^. Additionally, the lack of influence of socioeconomic status and other demographic factors on attrition in reviewed reports^27–30^ in comparison to other studies^20,37^ suggest that within minority populations of varying economic status, other factors must be considered. Findings that logistical barriers and unmet needs significantly affected program participation was consistent with previous literature^18,21^.

Our findings may not be generalizable as some results may be specific to the lifestyle modification programs investigated. Although the search strategy and study design were peer reviewed, only one team member performed screening, data abstraction, and quality review. Another limitation of this review was that all studies were based in the United States. While racial disparities exist in other countries, all of those reviewed here were specific to the US and the results may not be widely applicable outside the US.

## Conclusion

Efforts to reduce pediatric obesity among minority populations through the implementation of lifestyle change programs in their geographic area are still plagued by high rates of attrition. More research must be done to understand the complex needs of these communities in which systemic racism simultaneously drives the disparity in their obesity rates and presents obstacles to their treatment. Engagement with members of minority communities who have previously withdrawn from pediatric weight management programs can provide insight on how better programs can be designed. Including parents on program development teams; considering cultural influences on participation; and working with other stakeholders to reduce cost and other logistical barriers all have the potential to improve health outcomes for minority families.

## Data Availability

All data reviewed in this present work are available at:
1. doi:10.1007/s10995-020-03058-3
2. doi:10.1007/s10880-006-9015-x
3. doi:10.1016/j.orcp.2016.08.002
4. doi:10.3390/ijerph15071482
5. doi:10.1097/DBP.0b013e3181f17b1c

## Abbreviations

COVID-19: Coronavirus disease 2019
BMI: Body mass index
NHANES: National Health and Nutrition Examination Survey

## Acknowledgements

The author thanks Brian Piper, PhD for his mentorship and guidance as well as Michael Gionfriddo, Pharm.D, PhD for his feedback throughout the review process and peer review of search strategy and methods. The author also wishes to thank Tierney Lyons, MLS for assisting with the search and Mushfiq Tarafder, PhD, MPH, MBBS for his guidance.

## References

1. Wang Y, Lim H. The global childhood obesity epidemic and the association between socio-economic status and childhood obesity. Int Rev Psychiatry. 2012;24(3):176–188. doi:10.3109/09540261.2012.688195

2. Dai H, Alsalhe TA, Chalghaf N, Riccò M, Bragazzi NL, Wu J. The global burden of disease attributable to high body mass index in 195 countries and territories, 1990–2017: An analysis of the Global Burden of Disease Study. PLoS Med. 2020;17(7):e1003198. doi:10.1371/journal.pmed.1003198

3. Hemmingsson E. Early childhood obesity risk factors: socioeconomic adversity, family dysfunction, offspring distress, and junk food self-medication. Curr Obes Rep. 2018;7(2):204–209. doi:10.1007/s13679-018-0310-2

4. Chooi YC, Ding C, Magkos F. The epidemiology of obesity. Metabolism. 2019;92:6–10. doi:10.1016/j.metabol.2018.09.005

5. Stavridou A, Kapsali E, Panagouli E, et al. Obesity in children and adolescents during COVID-19 pandemic. Children. 2021;8(2):135. doi:10.3390/children8020135

6. Wu AJ, Aris IM, Hivert MF, et al. Association of changes in obesity prevalence with the COVID-19 pandemic in youth in Massachusetts. JAMA Pediatr. 2022;176(2):198–201. doi:10.1001/jamapediatrics.2021.5095

7. Obesity and overweight. Accessed February 5, 2022. https://www.who.int/news-room/fact-sheets/detail/obesity-and-overweight

8. Using the WHO growth charts | growth birth to 2 years | WHO | growth chart training | nutrition | DNPAO | CDC. Published January 23, 2019. Accessed February 5, 2022. https://www.cdc.gov/nccdphp/dnpao/growthcharts/who/using/index.htm

9. CDC. BMI for children and teens. Centers for Disease Control and Prevention. Published December 3, 2021. Accessed February 5, 2022. https://www.cdc.gov/obesity/childhood/defining.html

10. Fryar CD, Carroll MD, Afful J. Products - Health E Stats - prevalence of overweight, obesity, and severe obesity among children and adolescents aged 2–19 years: United States, 1963–1965 through 2017–2018. Published February 5, 2021. Accessed February 5, 2022. https://www.cdc.gov/nchs/data/hestat/obesity-child-17-18/obesity-child.htm

11. Caprio S, Daniels SR, Drewnowski A, et al. Influence of race, ethnicity, and culture on childhood obesity: implications for prevention and treatment. Diabetes Care. 2008;31(11):2211–2221. doi:10.2337/dc08-9024

12. Taveras EM, Gillman MW, Kleinman KP, Rich-Edwards JW, Rifas-Shiman SL. Reducing racial/ethnic disparities in childhood obesity: the role of early life risk factors. JAMA Pediatr. 2013;167(8):731–738. doi:10.1001/jamapediatrics.2013.85

13. Johnson VR, Acholonu NO, Dolan AC, Krishnan A, Wang EHC, Stanford FC. Racial disparities in obesity treatment among children and adolescents. Curr Obes Rep. 2021;10(3):342–350. doi:10.1007/s13679-021-00442-0

14. Mitchell N, Catenacci V, Wyatt HR, Hill JO. Obesity: overview of an epidemic. Psychiatr Clin North Am. 2011;34(4):717–732. doi:10.1016/j.psc.2011.08.005

15. Sahoo K, Sahoo B, Choudhury AK, Sofi NY, Kumar R, Bhadoria AS. Childhood obesity: causes and consequences. J Fam Med Prim Care. 2015;4(2):187–192. doi:10.4103/2249-4863.154628

16. Ward ZJ, Long MW, Resch SC, Giles CM, Cradock AL, Gortmaker SL. Simulation of growth trajectories of childhood obesity into adulthood. N Engl J Med. 2017;377(22):2145–2153. doi:10.1056/NEJMoa1703860

17. Kelleher E, Davoren MP, Harrington JM, Shiely F, Perry IJ, McHugh SM. Barriers and facilitators to initial and continued attendance at community-based lifestyle programmes among families of overweight and obese children: a systematic review. Obes Rev. 2017;18(2):183–194. doi:10.1111/obr.12478

18. Ball GDC, Sebastianski M, Wijesundera J, et al. Strategies to reduce attrition in managing paediatric obesity: A systematic review. Pediatr Obes. 2021;16(4):e12733. doi:10.1111/ijpo.12733

19. Spence ND, Skelton JA, Ball GDC. A proposed standardized approach to studying attrition in pediatric weight management. Obes Res Clin Pract. 2020;14(1):60–65. doi:10.1016/j.orcp.2019.11.004

20. Ligthart KAM, Buitendijk L, Koes BW, van Middelkoop M. The association between ethnicity, socioeconomic status and compliance to pediatric weight-management interventions – A systematic review. Obes Res Clin Pract. 2017;11(5):1–51. doi:10.1016/j.orcp.2016.04.001

21. Skelton JA, Beech BM. Attrition in paediatric weight management: a review of the literature and new directions. Obes Rev Off J Int Assoc Study Obes. 2011;12(501):e273–e281. doi:10.1111/j.1467-789X.2010.00803.x

22. Skelton JA, Irby MB, Geiger AM. A systematic review of satisfaction and pediatric obesity treatment: new avenues for addressing attrition. J Healthc Qual Off Publ Natl Assoc Healthc Qual. 2014;36(4):5–22. doi:10.1111/jhq.12003

23. Page MJ, McKenzie JE, Bossuyt PM, et al. The PRISMA 2020 statement: an updated guideline for reporting systematic reviews. BMJ. 2021;372:n71. doi:10.1136/bmj.n71

24. Data extraction forms. Accessed March 27, 2022. https://dplp.cochrane.org/data-extraction-forms

25. Risk of bias tools - RoB 2 for cluster-randomized trials. Accessed March 1, 2022. https://www.riskofbias.info/welcome/rob-2-0-tool/rob-2-for-cluster-randomized-trials

26. Risk of bias tools - ROBINS-I tool. Accessed March 1, 2022. https://www.riskofbias.info/welcome/home

27. Dhuper S, Bayoumi N, Dalvi J, Panzer B. The correlation between parental perceptions and readiness to change with participation in a pediatric obesity program serving a predominantly black urban community: a retrospective cohort study. Matern Child Health J. 2021;25(4):606–612. doi:10.1007/s10995-020-03058-3

28. Germann JN, Kirschenbaum DS, Rich BH. Use of an orientation session may help decrease attrition in a pediatric weight management program for low-income minority adolescents. J Clin Psychol Med Settings. 2006;13(2):169–179. doi:10.1007/s10880-006-9015-x

29. Kwitowski M, Bean MK, Mazzeo SE. An exploration of factors influencing attrition from a pediatric weight management intervention. Obes Res Clin Pract. 2017;11(2):233–240. doi:10.1016/j.orcp.2016.08.002

30. St. George SM, Petrova M, Kyoung Lee T, et al. Predictors of participant attendance patterns in a family-based intervention for overweight and obese Hispanic adolescents. Int J Environ Res Public Health. 2018;15(7):1482. doi:10.3390/ijerph15071482

31. Williams NA, Coday M, Somes G, Tylavsky FA, Richey PA, Hare M. Risk factors for poor attendance in a family-based pediatric obesity intervention program for young children. J Dev Behav Pediatr JDBP. 2010;31(9):705–712. doi:10.1097/DBP.0b013e3181f17b1c

32. Jelalian E, Hart CN, Mehlenbeck RS, et al. Predictors of attrition and weight loss in an adolescent weight control program. Obesity. 2008;16(6):1318–1323. doi:10.1038/oby.2008.51

33. Spence ND, Newton AS, Keaschuk RA, et al. Predictors of short- and long-term attrition from the parents as agents of change randomized controlled trial for managing pediatric obesity. J Pediatr Health Care Off Publ Natl Assoc Pediatr Nurse Assoc Pract. 2017;31(3):293–301. doi:10.1016/j.pedhc.2016.09.003

34. Rhodes ET, Boles RE, Chin K, et al. Expectations for treatment in pediatric weight management and relationship to attrition. Child Obes. 2017;13(2):120–127. doi:10.1089/chi.2016.0215

35. Sallinen Gaffka BJ, Frank M, Hampl S, Santos M, Rhodes ET. Parents and pediatric weight management attrition: experiences and recommendations. Child Obes. 2013;9(5):409–417. doi:10.1089/chi.2013.0069

36. Grow HMG, Hsu C, Liu LL, et al. Understanding family motivations and barriers to participation in community-based programs for overweight youth: one program model does not fit all. J Public Health Manag Pract JPHMP. 2013;19(4):E1–E10. doi:10.1097/PHH.0b013e31825ceaf9

37. Zeller M, Kirk S, Claytor R, et al. Predictors of attrition from a pediatric weight management program. J Pediatr. 2004;144(4):466–470. doi:10.1016/j.jpeds.2003.12.031

38. Barlow SE, Ohlemeyer CL. Parent reasons for nonreturn to a pediatric weight management program. Clin Pediatr (Phila). 2006;45(4):355–360. doi:10.1177/000992280604500408

