## Supplemental Table 1 for "Understanding Barriers toward Interventions for Childhood Obesity in Minority Communities: A Rapid Review"

### Supplemental Material

| Database | Search Strategy | Number of articles found | Date searched |
|---|---|---|---|
| PubMed | (“minority groups”[MeSH Terms] OR “minority group*”[tiab] OR “minority communit*”[tiab]) AND (“pediatric obesity”[MeSH Terms] OR “pediatric obesity”[tiab] OR “childhood obesity”[tiab]) AND ("Medically Underserved Area"[Mesh] OR "Race Factors"[Mesh] OR "Minority Health"[Mesh] OR "Healthcare Disparities"[Mesh] OR "Community Health Planning"[Mesh] OR "Health Services Accessibility"[Mesh]) | 11 | 1/18/22 |
| | (“minority groups”[MeSH Terms] OR “minority group*”[tiab] OR “minority communit*”[tiab]) AND (“pediatric obesity”[MeSH Terms] OR “pediatric obesity”[tiab] OR “childhood obesity”[tiab]) AND (Barriers[tiab] OR "Medically Underserved Area"[Mesh] OR Underserved[tiab] OR "Race Factors"[Mesh] OR "Minority Health"[Mesh] OR “Minority Health”[tiab] OR "Healthcare Disparities"[Mesh] OR Disparities[tiab] OR "Community Health Planning"[Mesh] OR "Community Health"[tiab] OR "Health Services Accessibility"[Mesh] OR Access*[tiab]) | 54 | 1/18/22 |
| | ("barriers"[tiab] OR "intervention"[tiab] OR "management"[tiab] OR "program*"[tiab]) AND ( "attrition"[tiab] OR "treatment refus*"[tiab] OR "treatment refusal"[Mesh]) AND ("child*"[tiab] OR "adolescen"[tiab] OR "pediatric"[tiab]) AND ("pediatric obesity"[Mesh] OR "pediatric obesity"[tiab] OR "overweight"[tiab]) AND ("minority health"[Mesh] OR "minorit*"[tiab] OR "medically underserved"[tiab] OR "disparities"[tiab] OR "race factors"[Mesh]) | 5 | 1/27/22 |

| | | | |
|---|---|---|---|
| | ("Young Adult"[Mesh] OR "Child"[Mesh] OR "Adolescent"[Mesh] OR "child*"[tiab]) AND ("Pediatric Obesity"[Mesh] OR "Overweight"[Mesh] OR "childhood obesity"[tiab] OR "Weight Loss"[Mesh] OR "Weight Reduction Programs"[Mesh]) AND ("Blacks"[Mesh] OR "Hispanic or Latino"[Mesh] OR "Ethnic and Racial Minorities"[Mesh] OR "minorit*"[tiab] OR "Black*"[tiab] OR "Hispanic or Latino"[tiab] OR "African American"[tiab]) AND ("Treatment Adherence and Compliance"[Mesh] OR "patient dropout"[tiab] OR "treatment refusal"[tiab] OR "attrition"[tiab] OR "treatment compliance"[tiab] OR "compliance"[tiab]) | 140 | 2/7/22 |
| | ( "Adolescen*"[tiab] OR "child*"[tiab]) AND ("Pediatric Obesity"[tiab] OR "Overweight"[tiab] OR "childhood obesity"[tiab] OR "Weight Reduction Programs"[tiab]) AND ("minorit*"[tiab] OR "Black*"[tiab] OR "Hispanic or Latino"[tiab] OR "African American"[tiab]) AND ("Treatment Adherence and Compliance"[tiab] OR "patient dropout"[tiab] OR "treatment refusal"[tiab] OR "attrition"[tiab] OR "compliance"[tiab]) | 18 | 2/7/22 |
| CENTRAL | (child* OR pediatric OR adolescen* OR teen) AND (obesity OR overweight OR "childhood obesity") AND ("weight management" OR "weight reduction" OR program*) AND (adherence OR attrition OR complian* OR retention OR "drop out" OR "patient engagement") AND (minorit* OR ethnic* OR race OR racial OR "minority group*" OR "minority communit*")<br><br>Restricted by: Title, Abstract, Keyword<br>All word variations searched | 89 | 2/3/22 |

| | | | |
|---|---|---|---|
| PsychINFO | ((pediatric obesity or childhood obesity or overweight) and (weight management or program) and (attrition or compliance or drop out or retention) and (minority or minority community or minority group or ethnicity or racial disparity or race)).ab. | 13 | 1/27/22 |
| | ((child or childhood or adolescence or pediatric or teen) and (obesity or overweight) and (weight management or program) and (adherence or attrition or compliance or drop out or retention) and (minority or minority community or minority group or ethnicity or racial disparity or race)).ab. | 10 | 1/27/22 |
| | (treatment compliance/ or compliance/ or treatment barriers/ or exp treatment dropouts/ or treatment refusal) AND (minority groups/ or alaska natives/ or american indians/ or blacks/ or hawaii natives/ or "latinos/latinas"/ or "race and ethnic discrimination"/ or "racial and ethnic groups" OR exp ethnic identity/ or sociocultural factors/ racial disparities/ or equity/ or health disparities OR "race (anthropological)"/) AND ( obesity/ or overweight/ or "obesity (attitudes toward)" OR weight loss) limit to (160 preschool age <age 2 to 5 yrs> or 180 school age <age 6 to 12 yrs> or 200 adolescence <age 13 to 17 yrs> or 320 young adulthood <age 18 to 29 yrs>) | 10 | 1/31/22 |

**Table 1.** Summary of search terms used in each database to identify relevant articles.
